# Mathematical Perspective of Covid-19 Pandemic: Disease Extinction Criteria in Deterministic and Stochastic Models

**DOI:** 10.1101/2020.10.12.20211201

**Authors:** Debadatta Adak, Abhijit Majumder, Nandadulal Bairagi

## Abstract

The world has been facing the biggest virological invasion in the form of Covid-19 pandemic since the beginning of the year 2020. In this paper, we consider a deterministic epidemic model of four compartments classified based on the health status of the populations of a given country to capture the disease progression. A stochastic extension of the deterministic model is further considered to capture the uncertainty or variation observed in the disease transmissibility. In the case of a deterministic system, the disease-free equilibrium will be globally asymptotically stable if the basic reproduction number is less than unity, otherwise, the disease persists. Using Lyapunov functional methods, we prove that the infected population of the stochastic system tends to zero exponentially almost surely if the basic reproduction number is less than unity. The stochastic system has no interior equilibrium, however, its asymptotic solution is shown to fluctuate around the endemic equilibrium of the deterministic system under some parametric restrictions, implying that the infection persists. A case study with the Covid-19 epidemic data of Spain is presented and various analytical results have been demonstrated. The epidemic curve in Spain clearly shows two waves of infection. The first wave was observed during March-April and the second wave started in the middle of July and not completed yet. A real-time basic reproduction number has been given to illustrate the epidemiological status of Spain throughout the study period. Estimated cumulative numbers of confirmed and death cases are 1,613,626 and 42,899, respectively, with case fatality rate 2.66 per cent till the deadly virus is eliminated from Spain.

## 1. Introduction

The world never thought that it would witness the worst catastrophe in the form of a pandemic viral disease when the first case of Covid -19 was detected in Wuhan, China, in December 20191 [1]. This highly infectious disease spread in 216 countries or territories within a short span of time, making it the largest pandemic in the history of infectious disease. This virus predominantly infects lungs as the spike protein (S-protein) on the cell surface of SARS-CoV-2 has high affinity to ACE2 (angiotensin converting enzyme 2) receptor, which is highly abundant on lung epithelial cells of human [2, 3]. This is the third attack of coronarus in the twenty-first century after the SARS CoV-1 in 2002-2003 and MERS in 2012. However, there are significant differences between the SARS CoV-2 infection and previous two covid infection in terms of total infected & death cases and terms of affected countries, though SARS-CoV-1 and SRAS-CoV-2 have more than 99% similarity in their genome sequence [4]. Understanding the severity of 2019-nCov, the World Health Organization declared a public health emergency on January 30, 2020, to alert the whole world and to get ready to fight against this highly infectious unusual pneumonia [5]. The WHO cry, however, was in vain and the virus spread in 114 countries with 118,000 positive cases and 4,291 casualties as of March 11. Assessing the “alarming levels of spread and severity, and by the alarming levels of inaction”, the Director General of WHO on March 11 announced Covid-19 outbreak as pandemic [6]. As of October 10, 2020, Covid-19 infection has spread in 216 countries/territories with cumulative confirmed cases more than 36 million and death cases over 1.05 million [1]. Covid-19 infection spreads from human-to-human through contact and inhaling the droplets containing the virus [5]. In absence of specific vaccine and drugs, human-to-human transmission of this virus can be reduced and prevented through non-pharmaceutical interventions (NPIs), which include using of musk, maintaining individual hygiene & safe distancing, lockdown, etc. [7, 8]. Such control measures can reduce the epidemic load by reducing the social mixing and can delay in achieving the epidemic peak [9, 10].

Mathematical model of infectious disease is central in epidemiology and might play an important role in forecasting the epidemiological burden of infectious disease [11]. A realistic model can predict the expected numbers of individuals to be infected and died during an epidemic [12]. It is also possible to predict time of attaining an epidemic peak and the expected cases at this peak. Such predictions help the healthcare providers and the government in resource planning and taking decision regarding control measures [13]. In the ongoing Covid-19 pandemic, countries like UK and USA have used mathematical models like a decision tool. For example, on the basis of mathematical model’s forecast of Imperial College, London, it was predicted that the UK health care system will be jeopardized with Covid-19 cases, and the country might have 500,000 deaths, and the UK Government after that put strict movement restriction [14]. The same epidemic model alerts USA by projecting that the country might face 2.2 million deaths if no controlling measures are imposed. Based on this mathematical result, USA implemented new guidelines for maintaining social distance [14].

Lots of mathematical models have been developed to give early stage epidemic predictions for the ongoing Covid-19 pandemic ([9, 10], [15]-[24]). All these models are deterministic type and do not consider uncertainty and variations in the parameters though it is obvious in the case of a growing epidemic. In particular, it has been shown that uncertainty is certain in the disease transmission rate of Covid-19 and there are large variation in its range [25]. Taking this into account, some stochastic models have been proposed for Covid-19 epidemic to address various epidemiological issues based on the simulation results [26, 27, 28, 29, 30]. In this work, we consider a deterministic SLIR (susceptible → latent → infected → recovered) epidemic model to encapsulate the Covid-19 epidemic of a given region and then extend this deterministic model to incorporate the stochasticity through parameter perturbation technique. Analysing both the deterministic and stochastic models, we give the disease extinction and persistence condition with respect to the basic reproduction number. We also verify our analytical results by performing a case study.

The remaining portion of this paper is arranged in the following sequence. The immediate next section describes the deterministic and stochastic model for Covid-19. Analysis of the deterministic model is presented in Section 3 and the same for the stochastic system is presented in the next section. A case study is performed with the Covid-19 epidemic data of Spain in Section 5. The paper ends with a discussion in Section 6.

## 2. Mathematical model

To construct a minimal model for Covid-19 disease transmission model, we divided the total population at any time *t, N*(*t*), of a geographical region into four compartments depending on the disease status of its population. All population, irrespective of age and sex, are assumed to be members of the susceptible class *S*. Susceptible population after coming into effective contact of infectious individuals *I* join the latent class *L*, whose individuals carry the virus but not infectious. Individuals of latent class on an average spent 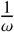 units of time in *L* class and then join infectious class *I*. Individuals of *I* class remain infectious for 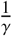 units of time and then either recovers at a rate γ or die due to covid infection at a rate *µ*. Thus, at any arbitrary time *t, N*(*t*) = *S* (*t*) + *L*(*t*) + *I*(*t*) + *R*(*t*). We further assume a death class *D* to represent the disease related death. The reason for considering this class is that death data is available for Covid-19 pandemic and can be used to fit the model parameters. This rate equation can also be used in the prediction of case fatality. If Λ be the constant input in susceptible class and *δ* is the natural death rate then the growth equation of each compartment can be represented by the following coupled nonlinear differential equations:

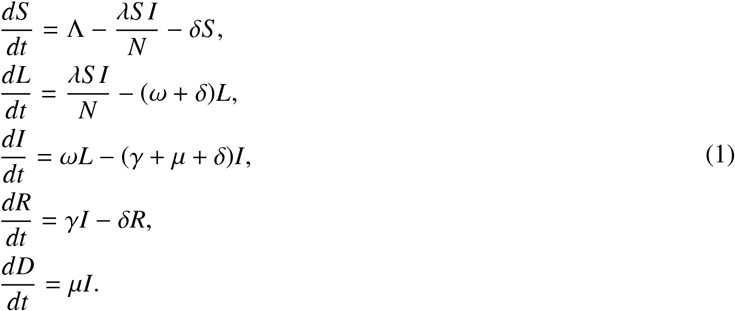

A very popular and effective technique to incorporate stochasticity into a deterministic model is parametric perturbation method [31, 32, 33, 34, 35]. Following parametric perturbation, the force of infection, *λ*, is replaced by

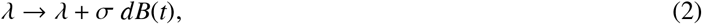

where *B*(*t*) is a standard independent Brownian motion and *σ* is the intensity of the noise. Thus, we extend the deterministic model (1) by incorporating white noise in the disease transmission term *λ* as

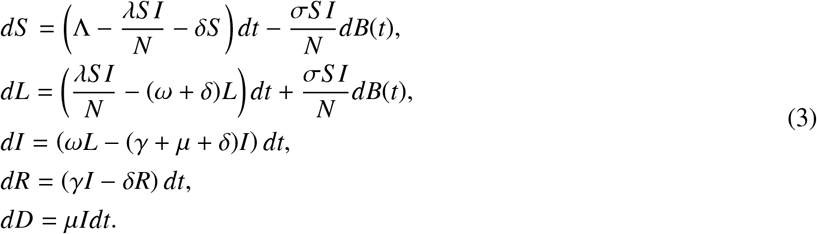

It is worth mentioning that the system (1) and (3) will be identical when *σ* = 0. Both the systems (1) and (3) will be analyzed with the initial conditions

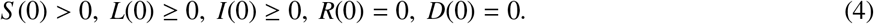

In the following, we always consider that the standard Brownian motion is defined on a complete probability space (*Ω, F*; *P*) with a filtration 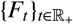 satisfying the usual conditions (i.e., it is right continuous and increasing while *F*_*0*_ contains all *P*-null sets).

## 3. Analysis of the deterministic model

From the biological point of view, we first show that the solutions of the system (1) exist uniquely, remain positive and bounded whenever starts with positive initial values.

### Proposition 3.1

*For the initial conditions specified in (4), solutions of the system (1) are nonnegative and uniformly bounded in*

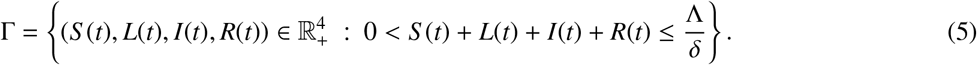

*Proof*. By adding the equations of system (1), we get

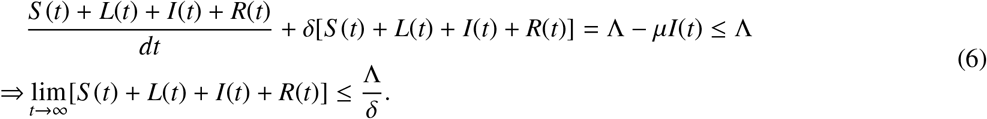

Hence all the solutions of the system (1) are ultimately bounded in the region Γ. The following lemma due to Nagumo [36] will be used to show that all solutions of the system (1) with initial conditions (4) are positive for all *t* ≥ 0.

### Lemma 3.1

*Consider a system* 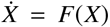, *where F*(*X*) = [*F*_1_(*X*), *F*_2_(*X*), …, *F*_*n*_(*X*)], *X* ∈ ℝ^*n*^, *with initial condition* 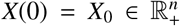. *If for X*_*i*_ = 0, *i* = 1, 2, …, *n*, 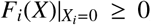, *then any solution of* 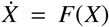 *with given initial condition, say, X*(*t*) = *X*(*t*; *X*_0_) *will be positive, i.e*., 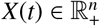.

Define *X*(*t*) = (*S* (*t*), *L*(*t*), *I*(*t*), *R*(*t*)). It is straight forward to show that

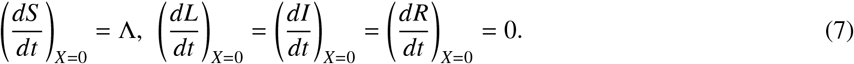

Lemma 3.1 then gives that all solutions of the system (1) starting with the initial conditions (4) are positive. Again,

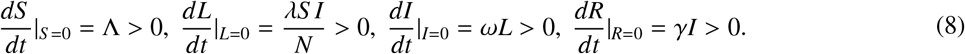

Thus, following [37], Γ is an invariant set of 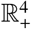. Therefore, all solutions of the system (1) with initial conditions (4) are positively invariant and ultimately bounded in Γ. □

### 3.1. Basic reproduction number

We determine the basic reproduction number (BRN), *R*_0_, associated with the deterministic system (1) using the “next generation matrix” approach [38, 39]. The infection subsystem of system (1) that describes the production of new infections and changes in the states capable of creating new infections in a completely susceptible scenario is given by

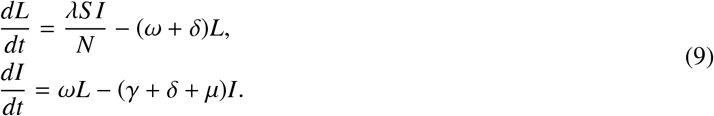

The transmission matrix and transition matrix associated with this infection subsystem (9) are, respectively, given by **T** and **Σ**, where

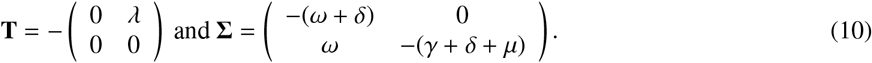

Then the basic reproduction number is given by

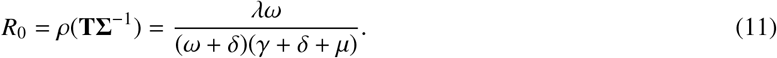

### 3.2. Equilibrium points

The system (1) has two equilibrium points, namely the disease-free equilibrium 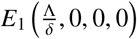, which always exists and the endemic equilibrium denoted by *E*^*^(*S* ^*^, *L*^*^, *I*^*^, *R*^*^). Define

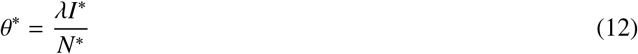

so that the equilibrium population densities of the system (1) at at *E*^*^ can be expressed as

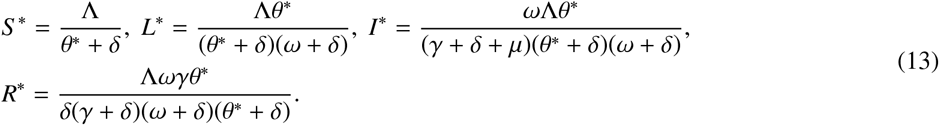

Then (12) gives

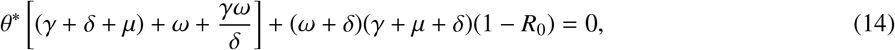

where *R*_0_ is given by (11). Clearly, we would get a unique positive θ^*^ and eventually a unique interior equilibrium *E*^*^ of the system (1) if *R*_0_ > 1.

In the following, we prove the stability results of different equilibrium points.

#### 3.2.1. Stability analysis

##### Theorem 3.1

*The disease-free equilibrium E*_1_ *is globally asymptotically stable if R*_0_ ≤ 1.

*Proof*. Consider the Lyapunov function given by

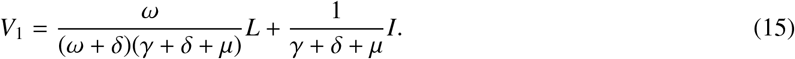

Differentiating *V*_1_ along the solutions of (1), we have

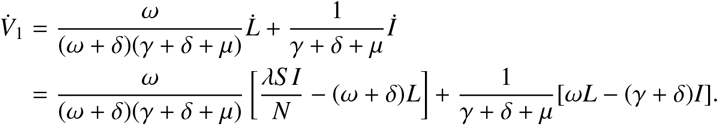

Noting that *S* (*t*) ≤ *S* (*t*) + *L*(*t*) + *I*(*t*) + *R*(*t*) = *N*(*t*) for all *t* ≥ 0,

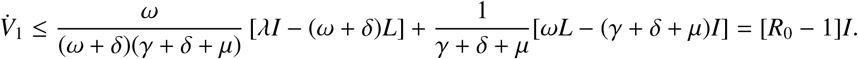

Thus, 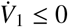 if *R*_0_ ≤ 1 with equality occurring at the disease free equilibrium *E*_1_. Therefore, using LaSalle’s invariance principle [40], one obtains (*L*(*t*), *I*(*t*)) → 0 as *t* → ∞. It gives that lim sup_*t*→∞_ *I*(*t*) = 0. Therefore, for any sufficiently small *ϵ* > 0, there exists a positive constant *M* > 0 such that lim sup_*t*→∞_ *I*(*t*) ≤ *ϵ* for all *t* > *M*. From (1), one can have

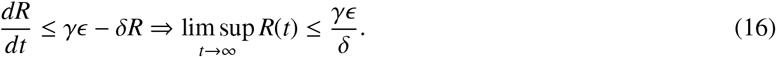

Letting *ϵ* → 0, we obtain lim sup_*t*→∞_ *R*(*t*) ≤ 0. Again, using the fact that lim inf_*t*→∞_ *I*(*t*) = 0, one can have lim inf_*t*→∞_ *R*(*t*) ≥ 0. Thus, we get lim_*t*→∞_ *R*(*t*) = 0. In a similar manner, one can show that 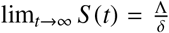. Therefore, all solutions of the system (1) with initial conditions in Γ eventually converge to the disease-free equilibrium *E*_1_ if *R*_0_ ≤ 1. Hence the theorem is proven. □

##### Theorem 3.2

*If exists, then the endemic equilibrium E*^*^ *is globally asymptotically stable*.

*Proof*. Define *κ*_*1*_ *= ω* + *δ, κ*_*2*_ *= γ* + *δ* + *µ* and consider the following Lyapunov function

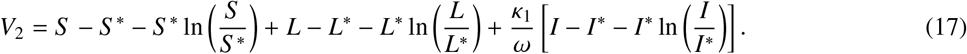

Differentiating (17) along the solutions of (1), we have

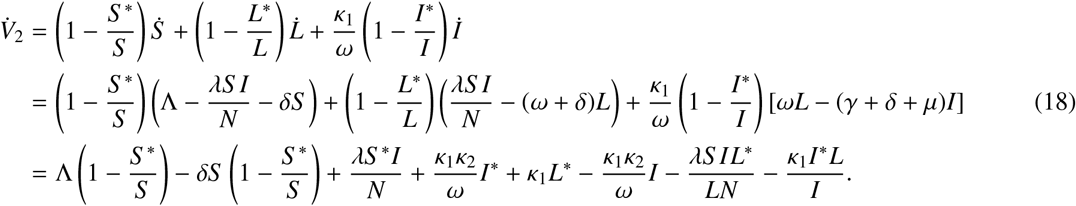

At *E*^*^, one have 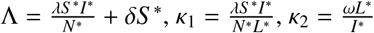. The above expression then becomes

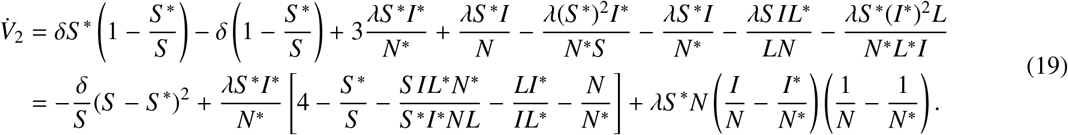

Using the fact that *A.M* ≥ *G.M*, one can write 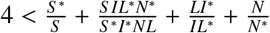. Define

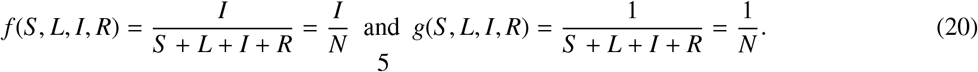

Thus, 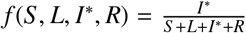. It should be noted that whenevr *I* = *I*^*^ holds then *S* = *S* ^*^, *L* = *L*^*^ and *R* = *R*^*^, giving rise to *N* = *N*^*^. Therefore, *f* (*S, L, I*^*^, *R*) = *f* (*S* ^*^, *L*^*^, *I*^*^, *R*^*^). Similarly, *g*(*S, L, I*^*^, *R*) = *g*(*S* ^*^, *L*^*^, *I*^*^, *R*^*^). Thus, from (19), we get

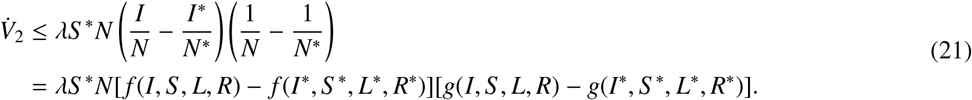

Clearly, *f* is monotone increasing in *I*, but *g* is monotone decreasing in *I*. So, if *I* > *I*^*^ then [*f* (*I, S, L, R*) − *f* (*I*^*^, *S* ^*^, *L*^*^, *R*^*^)] > 0 and [*g*(*I, S, L, R*)−*g*(*I*^*^, *S* ^*^, *L*^*^, *R*^*^)] < 0. However, if *I* < *I*^*^ then [*f* (*I, S, L, R*)− *f* (*I*^*^, *S* ^*^, *L*^*^, *R*^*^)] < 0 and [*g*(*I, S, L, R*) − *g*(*I*^*^, *S* ^*^, *L*^*^, *R*^*^)] > 0. In any case, 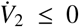. Moreover, the equality occurs when *S* = *S* ^*^, *L* = *L*^*^, *I* = *I*^*^. Therefore, by LaSalle’s invariance principle [40], (*S* (*t*), *L*(*t*), *I*(*t*)) → (*S* ^*^, *L*^*^, *I*^*^) as *t* → ∞. Hence lim sup_*t*→∞_ *I*(*t*) = *I*^*^. Thus, for sufficiently small *ϵ* > 0, there exists a positive constant *M*_1_ > 0 such that lim sup_*t*→∞_ *I*(*t*) ≤ *I*^*^ + *ϵ* for all *t* > *M*_1_. Then, from the fourth equation of (1), we get

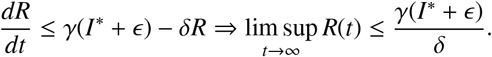

For *ϵ* → 0, we then have 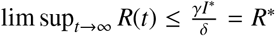. Similarly. using lim inf_*t*→∞_ *R*(*t*) ≥ *R*^*^, we obtain lim_*t*→∞_ *R*(*t*) = *R*^*^. It follows, whenever the endemic equilibrium *E*^*^ exists, all solutions of the system (1) with initial conditions in Γ converge to *E*^*^ as *t* → ∞. Hence the theorem is proven. □

##### Remark 3.1

It is to be mentioned that the considered system has two equilibrium points and stability of one hinders the stability of the other. As both the equilibrium points is globally stable, they must be locally stable under the same condition.

## 4. Analysis of the stochastic model

### Theorem 4.1

*For any initial values* (*S*(0), *L*(0), *I*(0), *R*(0)) 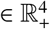, *there exists a unique global solution* (*S*(*t*), *L*(*t*), *I*(*t*), *R*(*t*)) 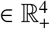 *of the system (3) for all t* ≥ 0 *and the solution will remain in* 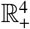 *with probability 1, i.e*., (*S*(*t*), *L*(*t*), *I*(*t*), *R*(*t*)) 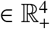 *for all t* ≥ 0 *almost surely (a.s)*.

*Proof*. Since the coefficients of the equations of system (3) are locally Lipscitz continuous for any initial value (*S*(0), *L*(0), *I*(0), *R*(0)) 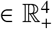, there is a unique local solution (*S*(*t*), *L*(*t*), *I*(*t*), *R*(*t*)) 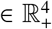 for all *t* ∈ [0, *τ*′), where *τ*′is the explosion time [48]. We now prove *τ* ′ = ∞ a.s., so that the solution becomes global.

Let *ν*_0_ > 0 be sufficiently large for every coordinate (*S*(0), *L*(0), *I*(0), *R*(0)) 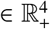 lying within the interval 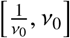. Now, for every integer *ν* > *ν*_0_, we define the stopping time

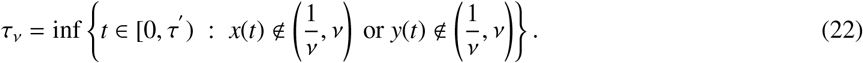

Here *τ*_*ν*_ is increasing as *ν* → ∞. Set lim_*ν*→∞_ *τ*_*ν*_ = *τ*_∞_, when *τ*_∞_ ≤ *τ* ′ a.s. We show that *τ*_∞_ = ∞ by the method of contradiction. Let us assume that our claim is not true and there exists two constants *T* > 0 and *ϵ* ∈ (0, 1) such that

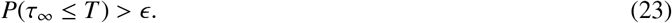

Thus, there exists an integer *ν*_1_ ≥ *ν*_0_ such that

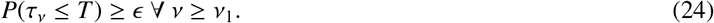

Noticing that *u* + 1 − ln *u* > 0 for all *u* > 0 and (*S*(0), *L*(0), *I*(0), *R*(0)) 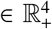, we define the following positive definite function

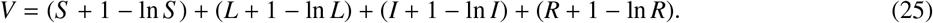

We consider *I*(*t*) ≥ 1 and *L*(*t*) ≥ 1 when the endemic equilibrium exists. Differentiating *V* using Ito’s formula [48], one can have

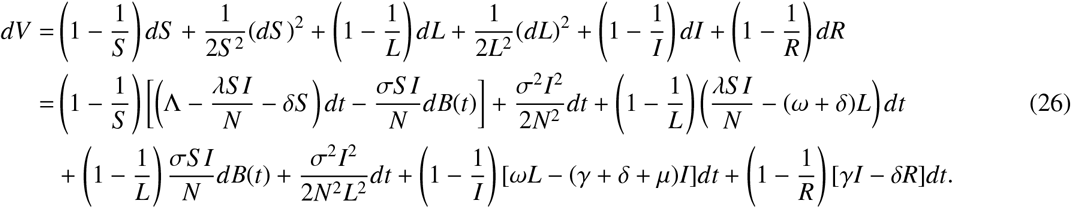

Observe that *u* ≤ 2(*u* + 1 − ln *u*) for all *u* > 0 and as *N* is the total population, we get *I*^2^ ≤ *N*^2^ and *I*^2^ ≤ *N*^2^*L*^2^. Hence, the above expression becomes

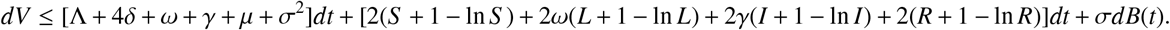

Let, Δ_1_ = Λ + 4*δ* + *ω* + *γ* + *µ* + *σ*^2^, Δ_2_ = max{2, 2*ω*, 2*γ*}, Δ_3_ = max{Δ_1_, Δ_2_}. Hence,

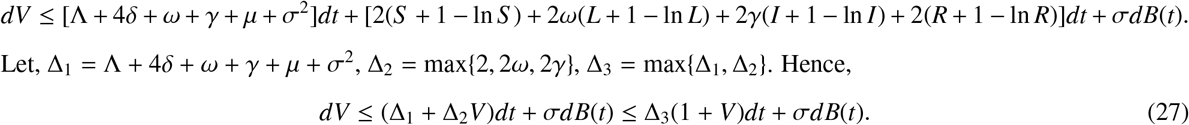

Integrating both sides of (27) from 0 to *t*_1_ ∧ *τ*_*ν*_ for any *t* ≤ *T* and taking expectation, we obtain

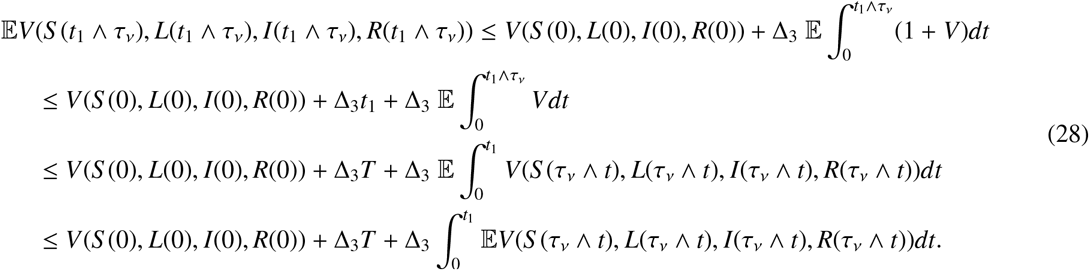

Now Gronwell’s inequality gives

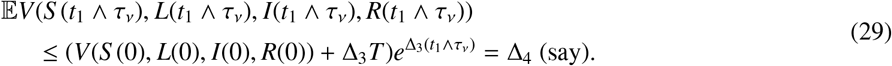

We set Λ_*ν*_ = {*τ*_*ν*_ ≤ *T*} for all *ν* ≥ *ν*_1_. Following (24), we then get *P*(Λ_*ν*_) ≥ *ϵ* for all *α* ∈ Λ_*ν*_. Clearly, at least one of *S* (*τ*_*ν*_, *α*), *L*(*τ*_*ν*_, *α*), *I*(*τ*_*ν*_, *α*), *R*(*τ*_*ν*_, *α*) is equal to either *ν* or 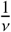. Hence *V*(*S* (*τ*_*ν*_), *L*(*τ*_*ν*_), *I*(*τ*_*ν*_), *R*(*τ*_*ν*_)) is no less than 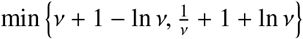. From (23) and (29), we obtain

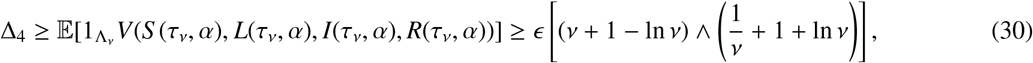

where 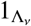 is the indicator function of Λ_*ν*_. Letting *ν* → ∞, one gets ∞ > Δ_4_ = ∞, a contradiction. Hence *τ*_∞_ = ∞ a.s. Hence the theorem is proven. □

### Lemma 4.1

*Let* (*S*(*t*), *L*(*t*), *I*(*t*), *R*(*t*)) 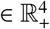 *be a solution of system (3) with positive initial value* (*S*(0), *L*(0), *I*(0), *R*(0)) 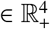. *Then*

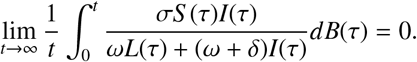

*Proof*. Let 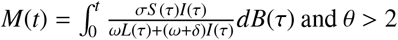 and *θ* > 2. Hence, by Burkholder-Davis-Gundy inequality [48] and Theorem 4.1, we get

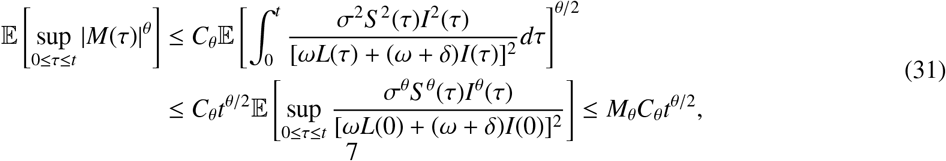

where 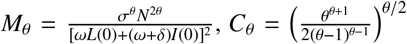. Then for any 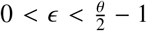, applying Doob’s martingle inequality [48] and using (31), we get

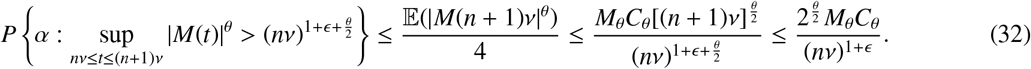

Thus, applying Borel-Cantelli lemma [48] on (32), for almost all *α* ∈ Λ, we obtain

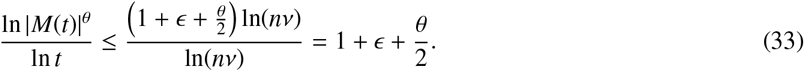

Hence, from (33), we have

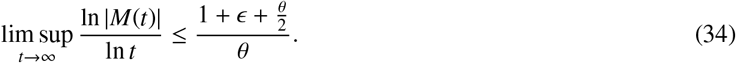

Letting *ϵ* → 0 in (34), one has

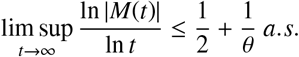

Then, for arbitrary positive constant 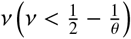, there exists a constant *K*(*α*) and a set Λ_*ν*_ such that *P*(Λ_*ν*_) ≥ 1 − *ν* and for *t* ≥ *K*(*α*), *α* ∈ Λ_*ν*_,

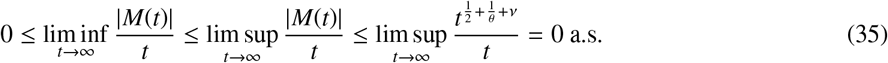

Therefore, following (35), one can get

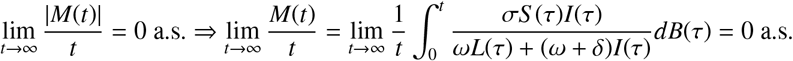

Hence the lemma is proven. □

### Remark 4.1

*As N*(*t*) *is the number representing the total human population, we consider N*(*t*) ≥ 1 *for all t* ≥ 0. *Hence*,

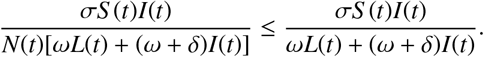

*Following Lemma 4.1, we have*

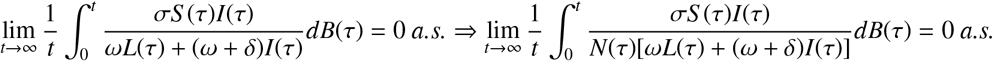

### Theorem 4.2

*The latent population L*(*t*) *and infected population I*(*t*) *of system (3) tends to zero exponentially almost surely if R*_0_ < 1, *where R*_0_ *is given by (11)*.

*Proof*. Let Let (*S*(*t*), *L*(*t*), *I*(*t*), *R*(*t*)) 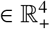 be a solution of the system (3) with positive initial value (*S*(0), *L*(0), *I*(0), *R*(0)) 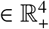. Define

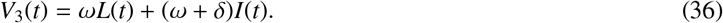

Differentiating (36) following Ito’s formula, one can get

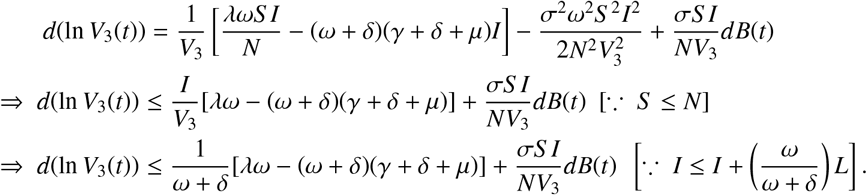

Integrating both sides of the last inequality from 0 to *t*, we get

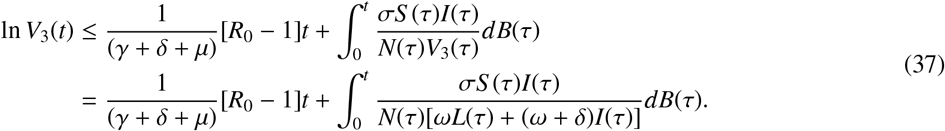

Taking limit superior as *t* → ∞ after dividing both sides of (37) by *t* and using Remark 4.1, we obtain

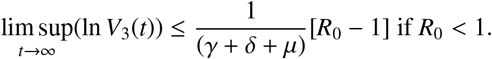

Then lim_*t*→∞_ *V*_3_(*t*) = lim_*t*→∞_[*ωL* + (*ω* + *δ*)*I*] = 0 a.s if *R*_0_ < 1. Again, as *ω* > 0, *ω* + *δ* > 0, we assert that lim_*t*→∞_[*ωL* + (*ω* + *δ*)*I*] = 0 ⇒ lim_*t*→∞_ *L*(*t*) = lim_*t*→∞_ *I*(*t*) = 0. Hence the result. □

### Theorem 4.3

*Let* (*S*(*t*), *L*(*t*), *I*(*t*), *R*(*t*)) 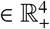 *be a solution of system (3) with positive initial value* (*S*(0), *L*(0), *I*(0), *R*(0)) 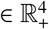. *If E*^*^(*S* ^*^, *L*^*^, *I*^*^, *R*^*^) *exists uniquely and* 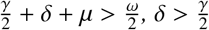 *are true then*

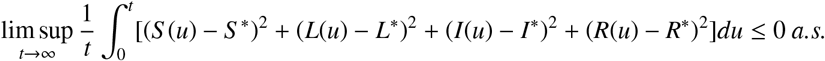

*Proof*. System (3) can be written as

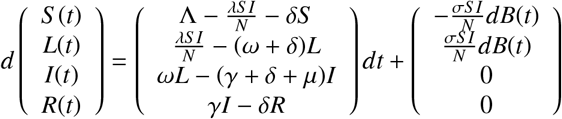

and the diffusion matrix is

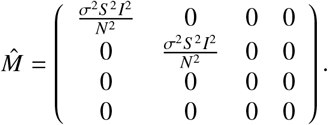

At *E*^*^, we have

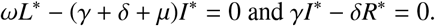

Define a *C*^2^ function 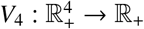 as

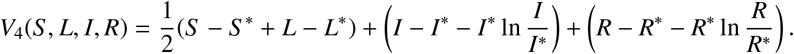

Applying Ito’s formula on *V*_4_, one gets

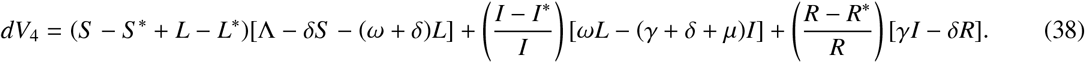

We consider *I*(*t*) ≥ 1 and *R*(*t*) ≥ 1 when the endemic equilibrium exists. Hence, from (38), we get

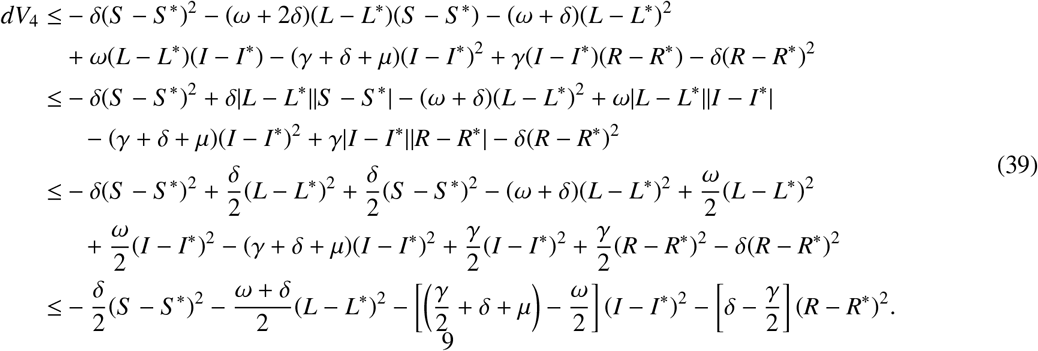

Under the restrictions 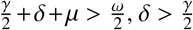 and defining 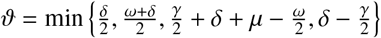, from the last inequality of (39), we get

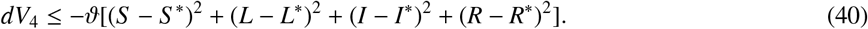

Integrating both sides of (40) from 0 to *t*, we have

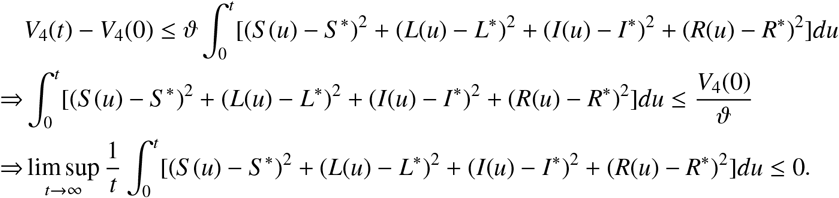

This completes the proof.□

## 5. Case study

Here we perform a case study considering the Covid-19 epidemic data of Spain, which witnessed the largest Covid-19 positive cases in Europe. The numbers of confirmed Covid-19 positive and death cases as of October 9, 2020, in Spain are 890,367 and 32,929, respectively, since the first case of Covid-19 was detected on January 31, 2020 [41]. The first phase lockdown with restricted movement was imposed in Spain on March 16, however, a more strict lockdown was came into force on March 30 [42] after observing 10,857 covid positive cases on a single day as of March 20 [43].

The data used in this study were taken from the online freely available repository Worldometer [43]. This study considers 228 days time series data of confirmed and death cases of Spain for the time span of February 25, when the Covid-19 positive cases were only 6 with zero death, to October 9, 2020, the completion date of the study period (both days included). The population of Spain as of January 1, 2020, was 47, 431, 256 [44] and this value was considered as the initial value of the susceptible population. The value of constant input into the susceptible class through birth (Λ) was considered as 1, 165 [45]. The average life expectancy of Spanish people is given to be 82.5 years [44]. Noting that 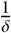 is the average life expectancy, the value of the parameter *δ* was then calculated as 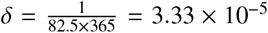. A Covid-19 infected individual remains in the latent class about 5.2 days [4], which gives rise to *ω* = 0.1924. The remaining three system parameters *λ, γ* and *μ* were estimated by fitting the model with the real data of infected and death cases in Spain.

Effect of different non-pharmaceutical controlling measures affects the parameter *λ*, the disease transmission efficiency or force of infection. The value of this parameter gradually decreases with the increasing length of lockdown, however, it increases with the withdrawal of lockdown. We, therefore, estimated *λ* in eight intervals each of 28 days (except the last one): (i) the first one from 25th February to 24th March, (ii) the second one from 25th March to 21st April, (iii) the third one from 22nd April to 19th May, (iv) the fourth one is from 20th May to 16th June, (v) the fifth one is from 17th June to 14th July, (vi) the sixth one is from 15th July to 11th August, (vii) the seventh one is from 12th August to 8th September and (viii) the final one is from 9th September to 9th October. We then used fminsearch optimization toolbox, SDE solver of Matlab and nonlinear least-square technique for the estimation of three parameters using our stochastic model (3). In Fig. 1, the actual data of infected and death cases were best fitted (*r*-squared values are 0.9994 and 0.9991) with *λ* = 0.542, 0.315, 0.192, 0.151, 0.164, 0.183, 0.271, 0.193 for the consecutive time intervals with *γ* = 0.095, *μ* = 0.11 and *σ* = 0.4. One can notice that the infection rate decreased during the lockdown period and then again increased due to its withdrawal. In fact, the daily positive cases were gradually declined after observing the maximum 961 cases on a single day on April 2. Then the second wave of covid infection in Spain started from the middle of July and peaked during the middle of September and reduced after that. Data fitting by the deterministic model (1) with the same parameter values is shown in Fig. 2.

**Figure 1:**
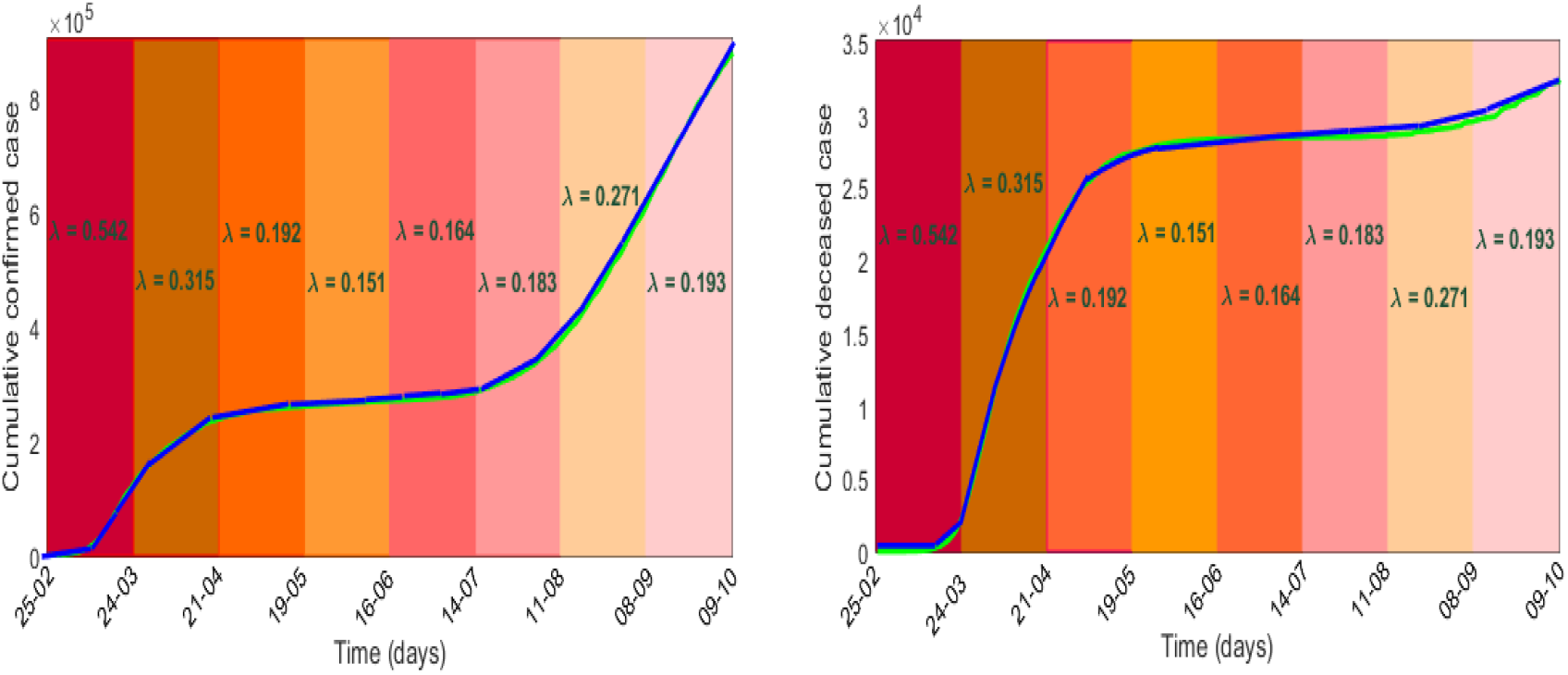
Stochastic model fitting: Simulated and actual values of the cumulative confirmed and death cases in Spain for the time span February 25 to October 9, 2020, are represented, respectively, by blue and green colour curves with *σ* = 0.4 and different force of infection, *λ*.

**Figure 2:**
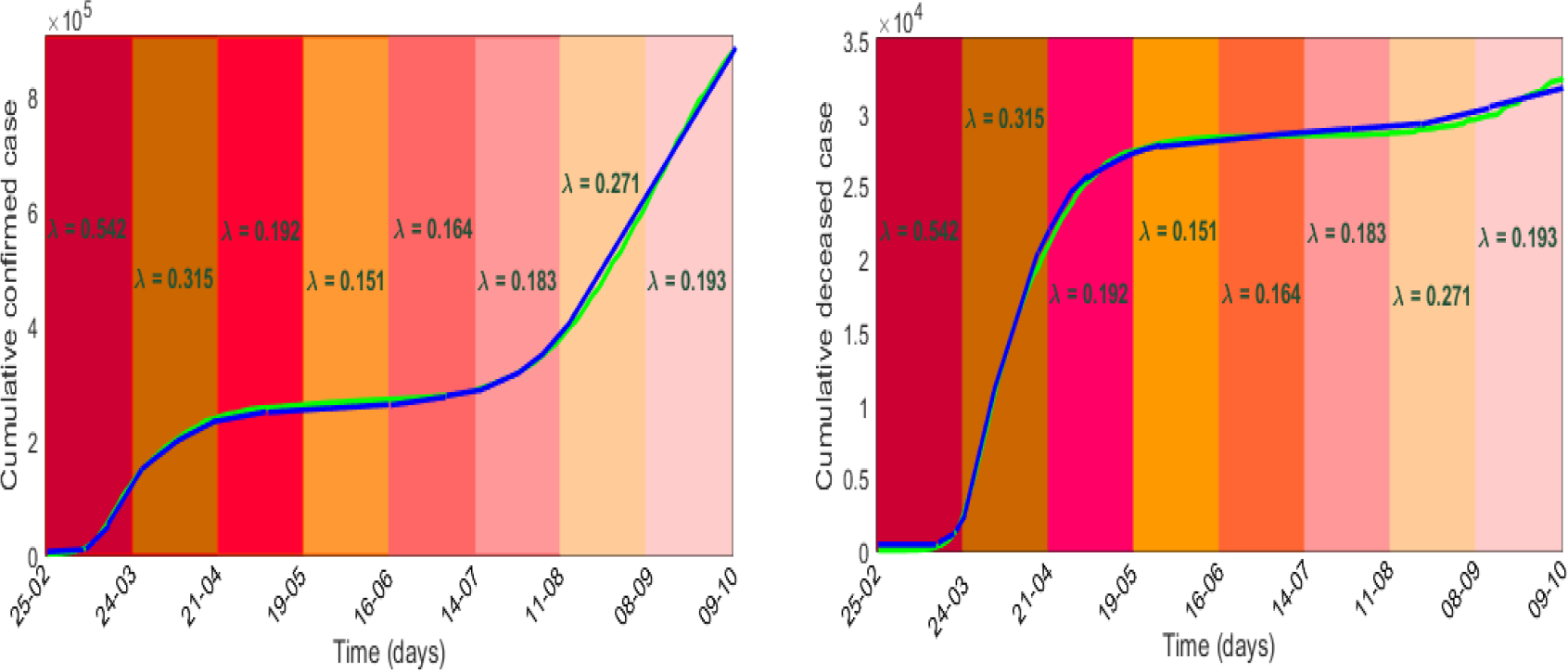
Deterministic model fitting: Simulated and actual values of the cumulative confirmed and death cases in Spain for the period February 25 to October 9, 2020, are represented, respectively, by blue and green colour curves.

Sensitivity analysis primarily identifies how the variation in the output of a system is affected by the variations of its input parameters. Applying the Latin Hypercube Sampling and Partial Rank Correlation Coefficient (LHS-PRCC) sensitivity analysis, we evaluated the PRCC values of the system parameters corresponding to each of the five variables, see Figure 3. Length of bars represents the effect of parameters on the system variables. It shows that the force of infection *λ* is the most sensitive one and has a strong effect on the growth of system populations with PRCCs significant to 0.00001 (*p*-values < 0.00001), further justifying our assumption of taking randomness in the parameter *λ*.

**Figure 3:**
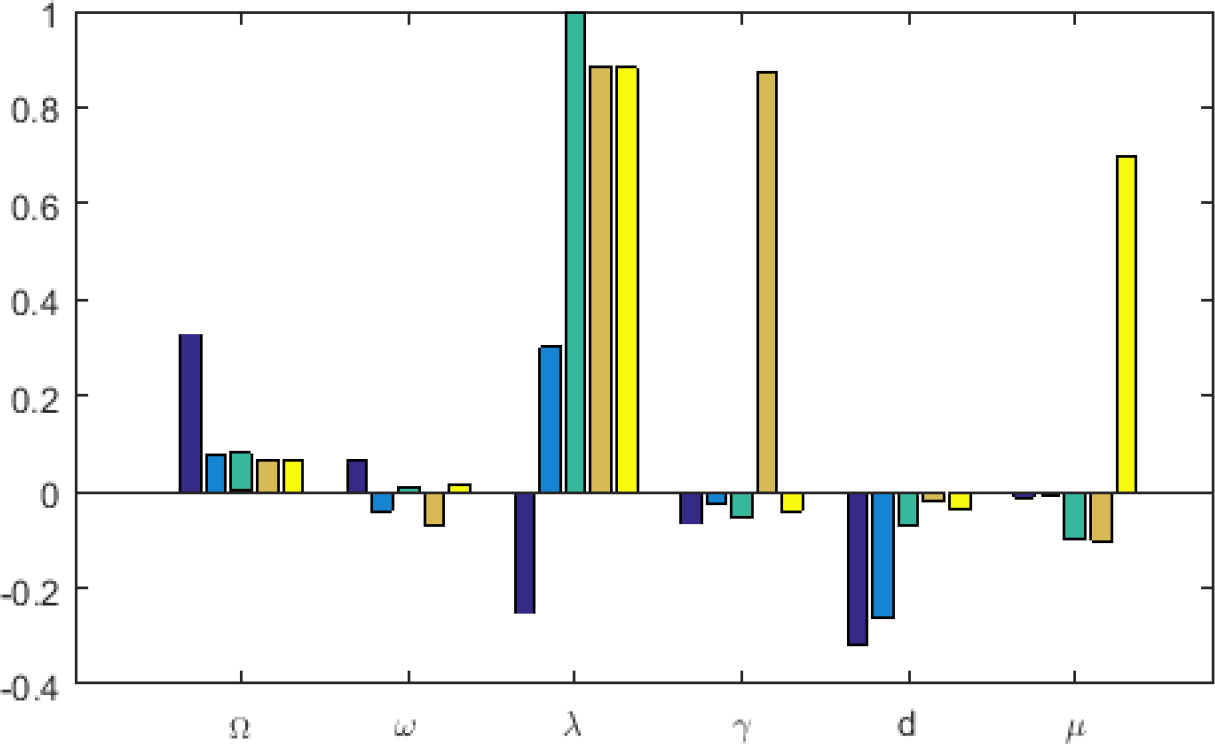
Global sensitivity analysis of the system parameters. Each parameter was varied over a range of ±2 fold from the baseline value. Partial ranked correlation coefficients (PRCC) sensitivity analysis (*p* < 0.00001) shows that the disease transmission coefficient, *λ*, is the most sensitive parameter. Baseline parameters are Λ = 1165, *ω* = 0.1924, *λ* = 0.54, *γ* = 0.095, *δ* = 3.37 × 10^−5^, *μ* = 0.11.

Basic reproduction number *R*_0_ is strongly dependent on the force of infection, while the number of infected individuals grows with increasing *R*_0_. In fact, the critical value *R*_0_ = 1 separates the endemic state (for *R*_0_ > 1) from the disease-free state (for *R*_0_ < 1). This has been demonstrated in Fig. 4 by the time series results. It shows that both the latent and infected class individuals of the deterministic system go to extinction for *R*_0_ < 1 (left upper panel) and persist for *R*_0_ > 1 (right upper panel). This result confirms the global stability of the two equilibrium points of the deterministic system with respect to the basic reproduction number, following the Theorems 3.1 and 3.2. Similar results for the stochastic system (3) are shown in the lower panel of Fig. 4.

**Figure 4:**
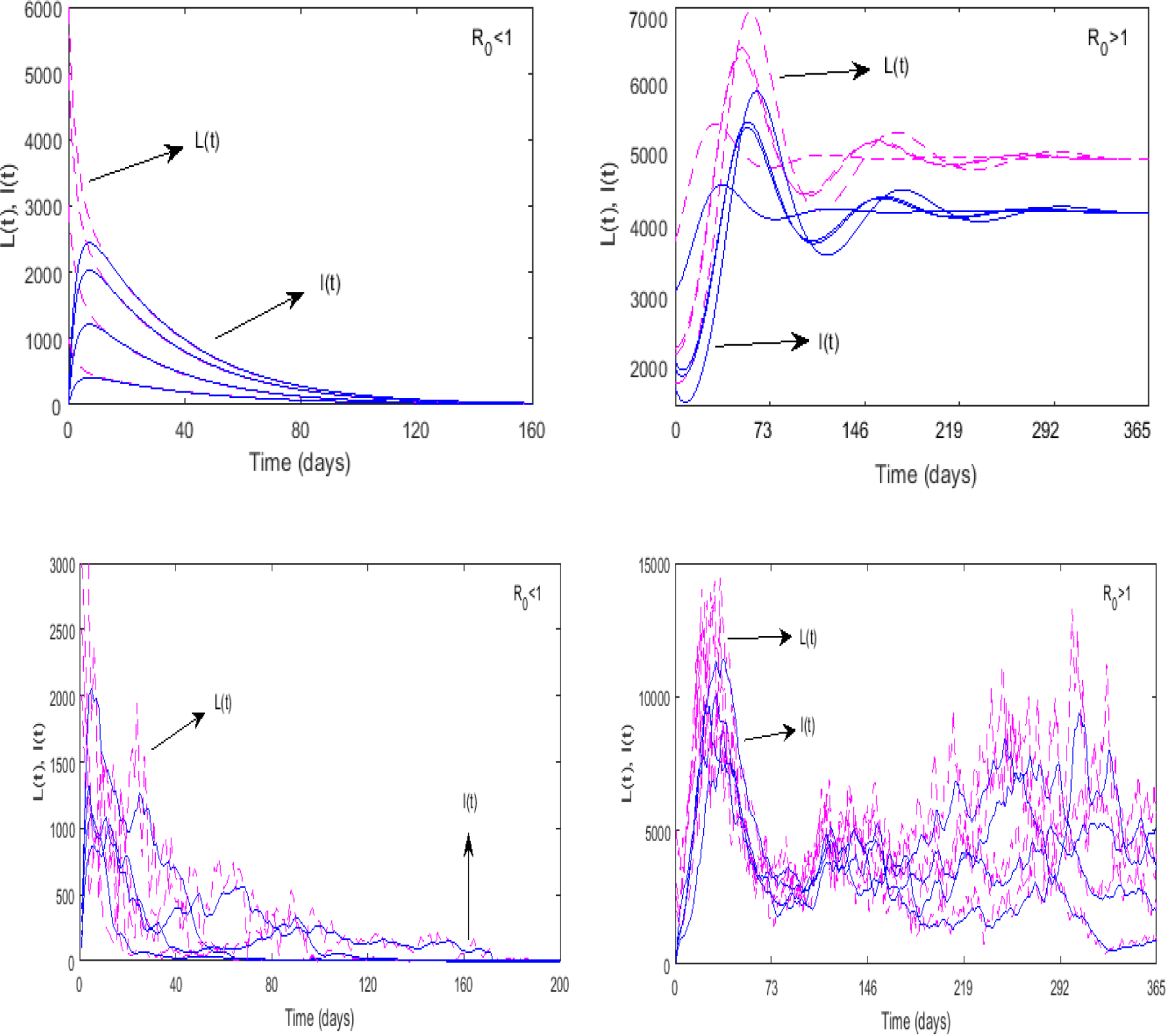
Disease extinction and persistence with respect to the basic reproduction number. Solutions of *L* and *I* classes of the deterministic system starting from different initial values go extinct (left panel) for *R*_0_ = 0.94 (< 1) and persist asymptotically (right panel) for *R*_0_ = 3.77 (> 1). The upper panel corresponds to the deterministic case and the lower panel for the stochastic case. Parameters are as in Figure 3 with *λ* = 0.18 for the left figure and *λ* = 0.74 for the right figure.

Real-time reproduction number (RTRN) is a data-driven analysis, which describes the epidemic status at every instant of time [46]. Considering the Covid-19 confirmed case data of Spain for the period March 3 to October 9, we plotted the real-time reproduction number, *R*_*t*_ (Fig. 5) with 95% confidence interval using the EpiEstim package of *R* software [47]. It is observed that RTRN goes below 1 on March 22 from its initial value 2.15 on March 3. After this date, the RTRN goes above one on September 1, however, it again goes below unity after 20th September. It is to be noticed that the daily cases during the second wave were maximum during the middle of September, which matches with the date of real-time reproduction number.

**Figure 5:**
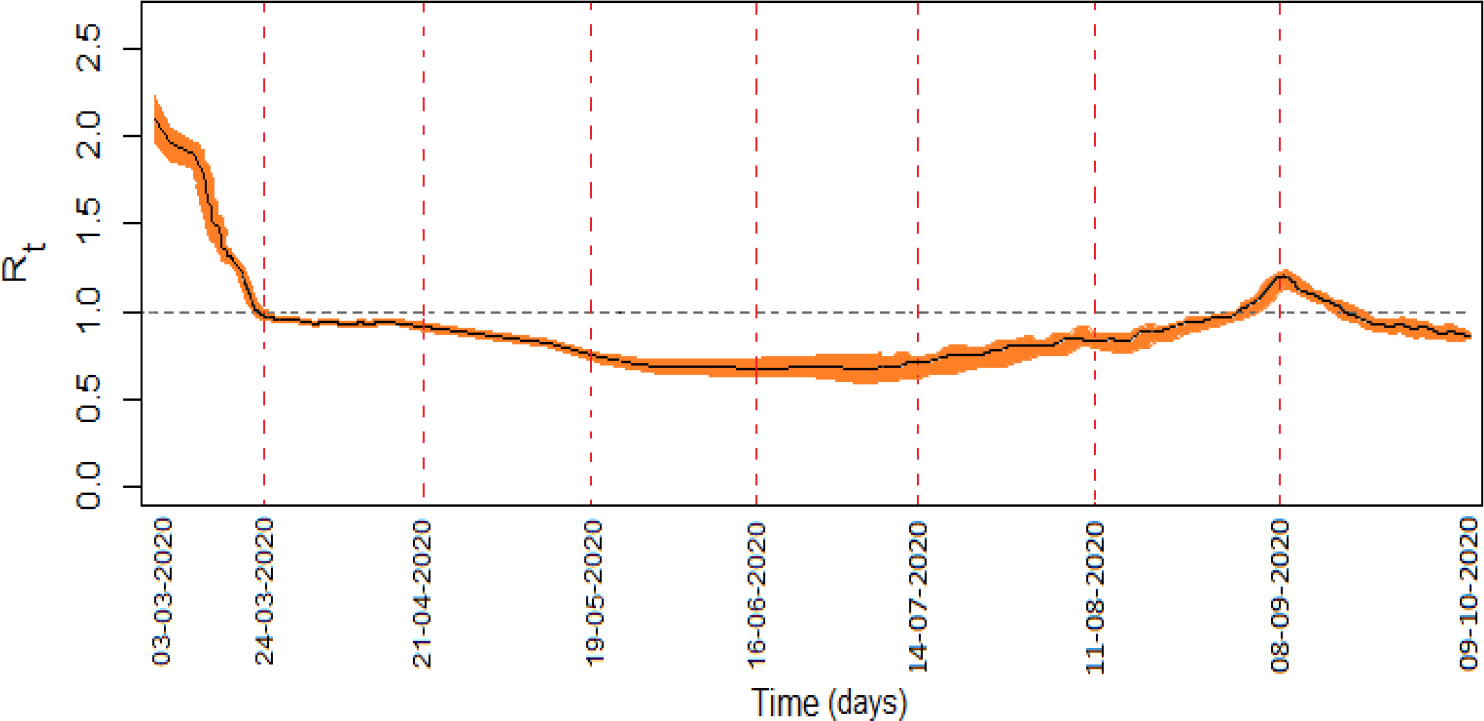
Real-time reproduction number (RTRN) of Spain for the period of March 3 to October 9 is represented by black line with 95% confidence interval. Parameters are as in Figure 3.

Though the stochastic system (3) originated from the deterministic system (1), the stochastic system has no interior equilibrium point but the later has. To compare the asymptotic behaviour of the solutions of the stochastic system (3) with that of the deterministic system (1), following Theorem 4.3, we plotted solutions of both the system in Fig. 6. It shows that solution of the stochastic system fluctuates around the deterministic solution, however, both solutions become identical as the noise intensity becomes too low.

**Figure 6:**
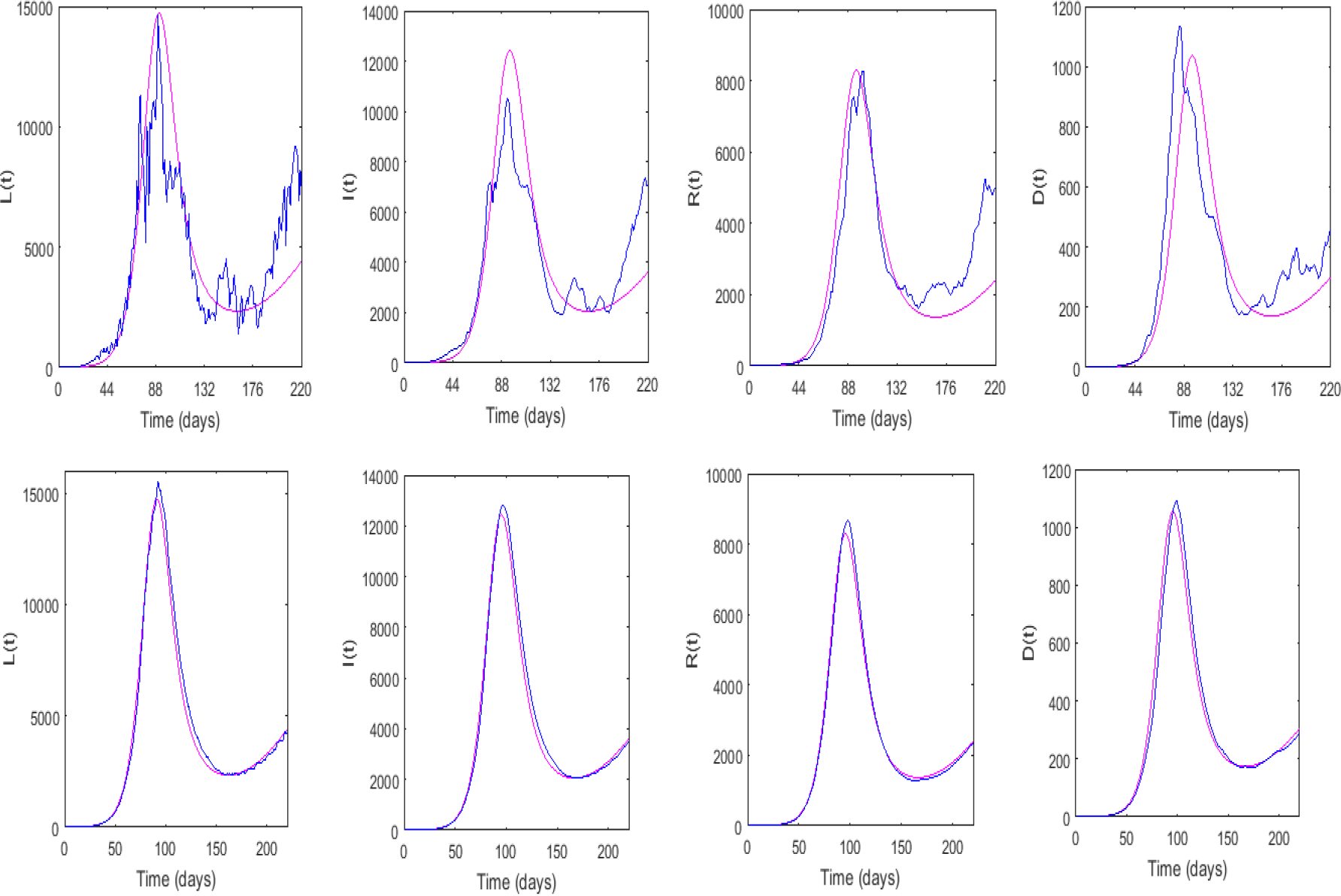
Upper panel: Asymptotic solutions of the deterministic and stochastic system populations with noise intensity *σ* = 0.4. Lower panel: Behaviour of the solutions for low value of the noise, *σ* = 0.01. Parameters are as in Figure 3.

In Fig. 7, we plotted per day actual data of confirmed cases with red colour and then plotted the simulation results in blue colour to predict the daily cases. It is interesting to note that Covid-19 infection was contained in Spain during May beginning to the first week of July. During this period the per day new cases was below 1,000. The situation, however, changed after the second week of July. The per day new case crossed 1,000 and steadily increased to 14,000 on September 18 by crossing 10,000 at the end of August. the epidemic curve again started to decline during the last week of September and reached to 6,000 per day on October 9 [43]. Fig. 7 thus shows two waves of infection.

**Figure 7:**
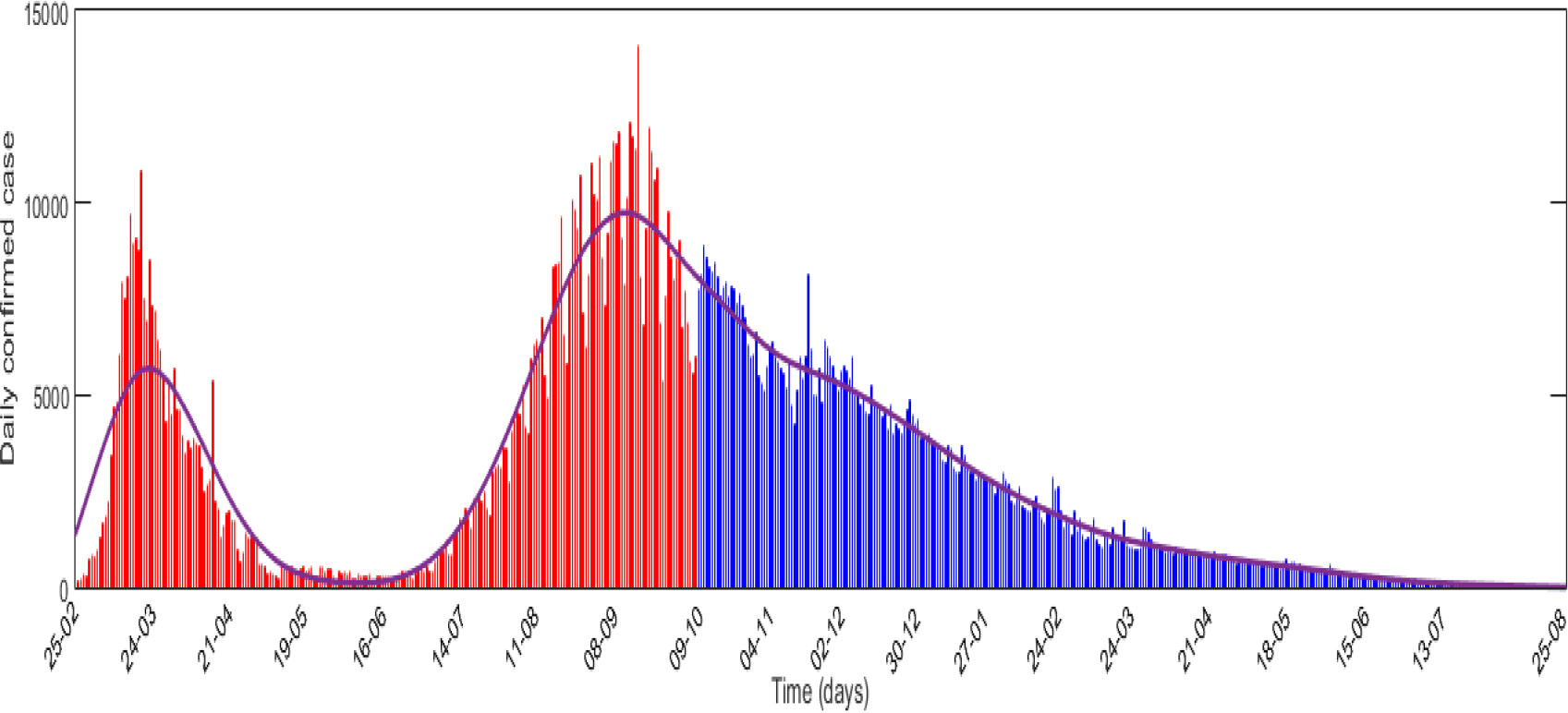
Daily cases in Spain. Actual values for the period February 25 to October 9 are represented by red bars. Predicted values for the period October 10, 2020, to August 25, 2021, are presented by blue bars. This figure clearly shows the second wave of Covid-19 infection during July-October after the first wave in March-April. The distribution curve is shown by the blue curve.

We also predicted the daily cases if the current train is maintained. This prediction was based on the stochastic model simulation results with *λ* = 0.193 and *σ* = 0.4. It shows that the epidemic curve has a Gaussian distribution with a long right tail. In the absence of any vaccine and if the present trend is maintained without further destruction, elimination of infection may be possible at the end of August 2021 (Fig. 8). The predicted number of confirmed and death cases on different dates are given in Table. 1. The case fatality rate, death per one hundred positive cases, is observed to vary from 3.41% to 2.88%.

**Figure 8:**
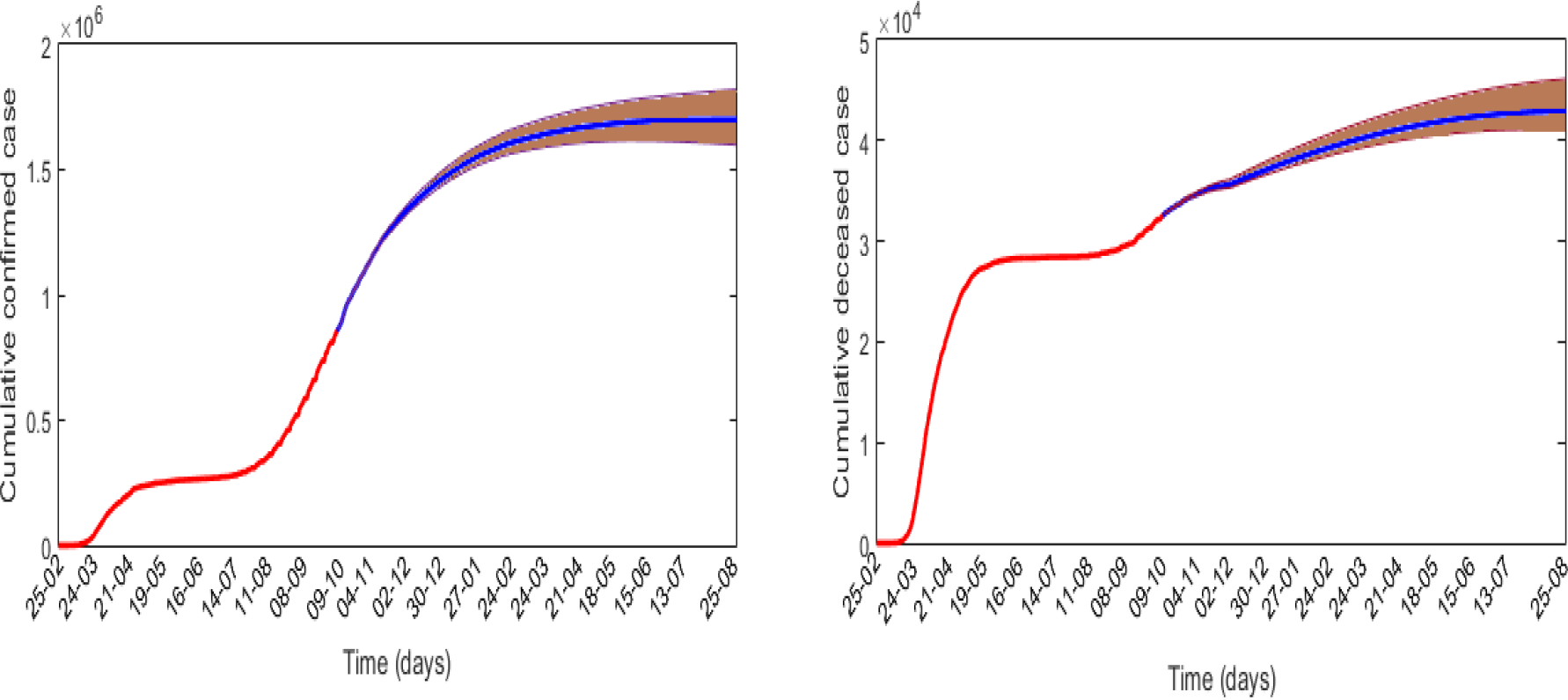
Cumulative confirmed and death cases in Spain from February 25 to October 9 are represented by magenta colour and the predicted values for the subsequent times are represented by blue colour with 95% confidence interval. It shows that the disease will be eliminated in the fourth week of August 2021, where the cumulative confirmed curve becomes flat.

**Table 1:** Estimated cumulative covid positive and death cases in Spain in the absence of vaccine and if the current trend is maintained

| Date | Confirmed cases | Death cases | CFR (%) |
| --- | --- | --- | --- |
| 23.10.2020 | 986,606 | 33,629 | 3.41 |
| 06.11.2020 | 1,078,178 | 34,622 | 3.21 |
| 20.11.2020 | 1,156,394 | 35,267 | 3.05 |
| 04.12.2020 | 1,237,538 | 35,667 | 2.88 |
| 25.08.2021 | 1,613,626 | 42,899 | 2.66 |

## 6. Discussion

Mathematical models are vital in infectious disease modelling. In epidemic models, the health status of the population of a given country is classified into different compartments and the flow of individuals from one compartment to other is then represented by a set of differential equations of deterministic or stochastic types. In the first type of epidemic models, all rate constants are assumed to be constant, though it is certain that the parameters, particularly the force of infection, are not constant but fluctuate around some mean value. Such fluctuations or randomness in the parameters may be successfully encapsulated by considering the stochastic extension of the deterministic model through parameter perturbation technique [48]. Acknowledging that both types of models have their own merits and demerits, it has been suggested that the results of the deterministic system should be verified with the results of the stochastic system. In this paper, we classified the entire population of a given geographical region into five compartments, viz., susceptible, latent, infectious and recovered classes, to represent the health status of its population infected by the coronavirus. A deterministic model was then proposed by considering the rate equations of each compartment. Different experimental and case studies have confirmed that there is a large variation in the infection rate of SARS-CoV-2 [25]. Furthermore, SARS-CoV-2 is a novel virus, whose aetiology is still not completely known. In such a case, it would be prudent to consider uncertainty in the infection rate. We, therefore, incorporated uncertainty in the disease transmission coefficient by adding the Gaussian white noise, assumed to be a good replicator of fluctuating phenomena [49]. Our PRCC analysis also showed that the force of infection was the most sensitive parameter of the system, giving further confirmation in adding noise to this parameter.

Both the deterministic and stochastic models were rigorously analyzed, and the disease eradication conditions were established. The basic reproduction number, which is assumed to be a milestone in every epidemic, was found to be the determinant of the epidemic fate. The basic reproduction number, computed from the next generation matrix of the deterministic system, was found to be sufficient to eliminate the infection if its value does not exceed 1, otherwise, the disease persists globally in the deterministic system. The same condition was found to be sufficient to eliminate the infection from the stochastic system (see Theorem 4.2), though the extinction time and nature of the solutions are quite different from its deterministic counterpart, they give the same result (see Fig, 4). The persistence result of the stochastic system significantly differs from the stochastic result. While the existing condition of the interior equilibrium is sufficient for the global disease persistency of the deterministic system, the stochastic system needs further conditions (see Theorem 4.3). Simulation results show that the solutions of the stochastic system fluctuate around the solution of the stochastic system, but the results become identical if the noise intensity is considered very small (see Fig. 6). In this sense, a stochastic solution is a generalization of the deterministic solution because one can obtain the deterministic solution from the stochastic solution but the converse is not true.

A case study with the Covid-19 epidemic data of Spain is performed by fitting our epidemic models. A real-time reproduction number (RTRN) has been presented to illustrate the epidemiological status of Spain on the daily basis. Covid-19 epidemic curve of Spain shows two waves of infection. The first wave was observed during March-April and the second wave was started at the middle of July and the waive is not completed yet. The epidemic peak during the first wave was observed on March 20 and the same was observed in the second wave after six months, September 18. The Covid-19 epidemic curve took a u-tern thereafter (see Fig. 7). The daily basis reproduction number curve also reciprocated the same feature of the epidemic situation. RTRN (real-time reproduction number) curve crossed the epidemic threshold value 1 twice, first time at the end of March and the second time on September 20 (Fig. 5). It indicates that the epidemic is in declined trained after these dates. RTRN as of October 9 is 0.86 and this value should be reduced close to zero for the elimination of infection. According to our prediction, if there is no vaccine and the system is not further perturbed, Spain will eliminate SARS-CoV-2 infection at the end of August 2021 with 1, 613, 626 infectives and 42, 899 deaths, giving rise to 2.88% case fatality. The Covid-19 situation in Spain indicates that infection may return if there is a lack of strong monitoring. Strong participation of common people in the containment program of coronavirus is a must, otherwise, multiple waves of infection may occur unless as there is no vaccine.

It is to be mentioned that we have kept our model simple, keeping only the most vital compartments required to represent the SARS-CoV-2 infection spreading mechanism. Although there are more complex models that include other epidemic compartments of coronavirus disease. This simple model, however, allowed us to determine analytically the disease extinction and persistence criteria for both the deterministic and stochastic systems. Secondly, we wanted to fit our models with the Covid-19 epidemic data of a country. The data in different repositories give only three classified data of confirmed, recovered and death cases. Thus, a higher compartmental model, which involves many additional parameters, will inherently accumulate more errors in the processes of estimation of the additional parameters, simply due to unavailability of data from which they can be estimated. The simple mathematical model has been used successfully in estimating infection horizon of Covid-19 with a data-driven approach [19]. Once more classified data are available, specifically the age-structured data, then more complex models would be helpful for in-depth study of Covid-19 pandemic. Despite its simplicity, the model can be used successfully in predicting the epidemiological burden of Covid-19 epidemic of any country.

## Data Availability

All data are available freely in the following depository Worldometer

https://www.worldometers.info/coronavirus/country/spain/

